# The Digital Analytic Patient Reviewer (DAPR) for COVID-19 Data Mart Validation

**DOI:** 10.1101/2021.05.30.21257945

**Authors:** Heekyong Park, Taowei David Wang, Nich Wattanasin, Victor M. Castro, Vivian Gainer, Sergey Goryachev, Shawn Murphy

## Abstract

**Objective:** To provide high-quality data for COVID-19 research, we validated COVID-19 clinical indicators and 22 associated computed phenotypes, which were derived by machine learning algorithms, in the Mass General Brigham (MGB) COVID-19 Data Mart.

**Materials and Methods:** Fifteen reviewers performed a manual chart review for 150 COVID-19 positive patients in the data mart. To support rapid chart review for a wide range of target data, we offered the Digital Analytic Patient Reviewer (DAPR). DAPR is a web-based chart review tool that integrates patient notes and provides note search functionalities and a patient-specific summary view linked with relevant notes. Within DAPR, we developed a COVID-19 validation task-oriented view and information extraction logic, enabled fast access to data, and considered privacy and security issues.

**Results:** The concepts for COVID-19 positive cohort, COVID-19 index date, COVID-19 related admission, and the admission date were shown to have high values in all evaluation metrics. For phenotypes, the overall specificities, PPVs, and NPVs were high. However, sensitivities were relatively low. Based on these results, we removed 3 phenotypes from our data mart. In the survey about using the tool, participants expressed positive attitudes towards using DAPR for chart review. They assessed the validation was easy and DAPR helped find relevant information. Some validation difficulties were also discussed.

**Discussion and Conclusion:** DAPR’s patient summary view accelerated the validation process. We are in the process of automating the workflow to use DAPR for chart reviews. Moreover, we will extend its use case to other domains.

## Introduction

### Background

When the COVID-19 pandemic arrived in the US [1], there was a growing demand for COVID-19-related data in the research community. Providing accurate and fluent data in a timely manner is essential to conquering this unprecedented disease. *Mass General Brigham (MGB) Research Information Science and Computing (RISC)* quickly created data tools, including the *COVID-19 Data Mart* and the *COVID-19 Summary Table* [2], and made available this information to research groups across the MGB system [3-12]. The COVID-19 Data Mart contains COVID-19-tested patients and their associated data, both structured and unstructured. It provides direct access to data tables as well as one-stop analysis options without having to pull data out of the Mart. The COVID-19 Summary Table holds COVID-19 positive patient data in discrete data columns. It is designed for quick identification and analysis of the COVID-19 positive patient cohort. By the time we performed this study in July 2020, the COVID-19 Data Mart reached over 88,000 patients and the COVID-19 Summary Table accumulated over 17,000 patients.

However, the advent of the new disease brought many challenges in providing high-quality data. In the beginning, we did not have a diagnosis code for COVID-19, and there were a lot of false negatives in COVID-19 test results. Even after the ICD-10 [13], LOINC [14], and CPT [15] codes for COVID-19 were released, we could not solely rely on the coded data to identify COVID-19 positive patients. First, most of the codes are recorded for billing purposes at the end of a hospitalization or after the patient is discharged. If a patient’s data is integrated into a data mart while the patient is still in hospital, code information is not yet available. Second, COVID-19 information can be miscoded due to the time gap between a treatment and a COVID-19 test result. For example, some patients were coded as COVID-19 patient initially but turned out to be negative later. Lastly, transferred patients often do not have a COVID-19 test result in our electronic health record (EHR) system. Instead, the information is only available in narrative reports, making it harder to categorize them. Therefore, various new algorithms are developed and applied to infer key information.

Associating COVID-19 data with clinically relevant information was also challenging. Since we did not fully understand COVID-19, it was hard to decide, for example, what are the comorbidities and what information would be helpful. Moreover, the influx of new patients created exceptional situations. We did not have data in our system if COVID-19 patients were transferred in. Large portions of them were healthy prior to admission so they had no rich data to mine. Large volume of missing data raises concerns about the reliability of our phenotyping algorithms [16-28]. In addition, during the surge, many seriously ill patients did not get coded as having an ICU visit (i.e., a major severity indicator) due to the bed shortage. Therefore, validating the COVID-19 data sets became an urgent goal.

### Problems

Unlike other validations, COVID-19 data validation needed to be completed in a short time, targeted broad disease domains, and was expected to require more note reviews. Our previous validation efforts [29-34] typically focused on a single target disease and involved a few experts on that disease to establish a gold standard by reviewing charts. However, the unprecedented urgency of the pandemic and the novelty of the disease meant that we needed to rely on volunteers with diverse clinical backgrounds and different chart review skills. The diversity in clinical background meant that some validation goals were more difficult for some reviewers and easier for others, depending on their clinical expertise. In addition, COVID-19 patients often lack reliably coded data, as many of them are new to our system, so our reviewers had to be even more reliant on text notes that describe patient history in natural language.

## Objective

Our aim was to validate data in the COVID-19 Data Mart to provide a high-quality data resource to the research community at Mass General Brigham. In the first validation phase, we validated COVID-19 information and 22 phenotypes of COVID-19 positive patients. The target data were derived facts computed by rule-based or machine learning algorithms. The task was reviewing patient history manually to verify the derived values. To support the above objectives, we built the *Digital Analytics Patient Reviewer (DAPR)* chart review tool. In this paper, we describe how we transformed DAPR to serve the COVID-19 Data Mart validation work, how we streamlined the validation process to utilize DAPR, the validation work itself, and the results.

## Materials and methods

### Data

We used the COVID-19 Summary Table to validate the MGB COVID-19 Data Mart. The COVID-19 Summary Table originates from the MGB COVID-19 Data Mart. It includes COVID-19 positive patient data, one row for every patient. The data types in the columns include patient demographics, EPIC Infection flags, COVID-19 PCR and antibody laboratory tests, inpatient admission information and phenotype data derived by various algorithms. We selected 150 patients to validate the MGB COVID-19 Data Mart. The patients were randomly chosen from the summary table patients who have at least 1 target phenotype in their history.

We asked the validators to validate the COVID-19 patient cohort indicator (Positive), index date of COVID-19 positive status, admission associated with COVID-19 (Y/N), COVID-19 admission date, and 22 machine learning phenotypes (Y/N) considered to be associated with COVID-19 (Table 1). Both Y and N values should be validated. We provided the data along with additional information: patient identifiers, demographics, COVID-19 status and tests results, COVID-19 severity indicator, and COVID-19 flags.

**Table 1.** Target data for the COVID-19 Data Mart validation

| Category | Target Data | Value<br>Format |
| --- | --- | --- |
| COVID-19 status / Tests | COVID-19 patient cohort | Positive |
| COVID-19 flags | COVID-19 index date | date |
| COVID-19 admissions | COVID-19 admission | Y/N |
|  | COVID-19 admission date | date |
| Disease phenotypes | Atrial fibrillation Curated Phenotype, Atrioventricular block<br>Curated Phenotype, Coronary atherosclerosis Curated<br>Phenotype, Crohns disease Curated Phenotype, Deep vein<br>thrombosis Curated Phenotype, Female infertility Curated<br>Phenotype, Heart valve disorders Curated Phenotype,<br>Hyperlipidemia Curated Phenotype, Hyperparathyroidism<br>Curated Phenotype, Hypertension Curated Phenotype,<br>Hypothyroidism Curated Phenotype, Myocardial infarction<br>Curated Phenotype, Obstructive sleep apnea Curated Phenotype,<br>Peripheral vascular disease Curated Phenotype, Polycystic<br>ovaries Curated Phenotype, Pulmonary heart disease Curated<br>Phenotype, Renal failure Curated Phenotype, Systemic lupus<br>erythematosus Curated Phenotype, Tobacco use disorder Curated<br>Phenotype, Type 2 diabetes Curated Phenotype, Type 2 diabetes<br>Curated Phenotype, and Ulcerative colitis Curated Phenotype | Y/N |

### Study Participants and the Center for COVID Innovation Working Group

In March 2020, MGB initiated the *Center for COVID Innovation* [35] to help develop innovations for the most pressing COVID-19 issues. In the *Clinical Trial Tools & MGB COVID-19 Data Mart Working Group*, a multidisciplinary group of researchers, including infectious disease specialists, physicians, biostatisticians, and informaticians, had weekly virtual meetings to discuss the COVID-19 Data Mart and its validation [36]. We introduced the needs for COVID-19 data validation to the working group and had discussions on determining important COVID-19 features and how to identify them in the EHR. Through this active communication, 14 participants volunteered for COVID-19 data validation. Moreover, this meeting led us to revive DAPR for chart review.

### Digital Analytic Patient Reviewer (DAPR)

DAPR is rooted from a decommissioned tool named the *Queriable Patient Inference Dossier (QPID)* [37], which provides a patient-specific summary view that displays medical concepts linked with relevant notes and allows users to search notes for clinical terms in a web-based client application. It integrates notes across different information systems and uses Natural Language Processing (NLP) to pull relevant information. The NLP rules are represented by QPID Query Language (QQL) and can be incorporated into a user interface component. If a user hovers over one of a problem in the view, it drills into the notes where the problem is described. When searching, it highlights the term in preview search results and in the actual note. The QPID was originally designed for clinical use in radiology department. However, due to its useful features for patient chart review, it had often been used for validation by other clinical and research groups. We reimplemented the service as a research tool for our COVID-19 data validation task.

First, we created a COVID-19 validation task-oriented summary view (Figure 1). We selected COVID-19-relevant information and past medical history that would help the validation and reorganized the summary list. We placed COVID-19-relevant concepts on the top row and past medical history to the bottom. In the ‘COVID-19’ row, we sectioned the category into COVID - 19 status, risk factors, severity, and management. For each section, we listed frequently used concepts in clinical settings. In the bottom row, we reused part of the summary list existed in the former version, since the list have been useful for various chart reviews. Furthermore, we integrated the 22 target phenotypes into this category as the ‘PHENOTYPES’ section. We added 127 new summary items and developed information extraction logic to find relevant notes and highlight key information. We built the logic in QQL and specified patterns using extended keywords and code information as well as date and note type constraints.

**Figure 1.**
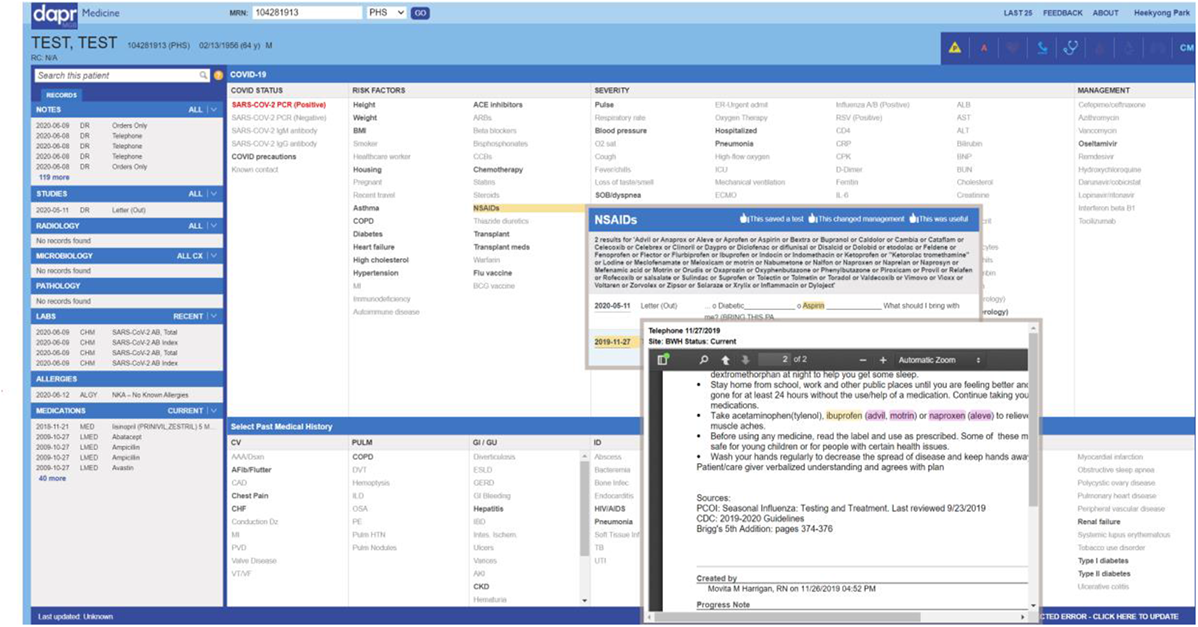
DAPR screen. DAPR is a patient specific chart review tool. If users enter a patient identifier on the top search box, it displays patient name, MGB site name, and demographic information on the top, all note list categorized by note types in the left panel, and a summary view generated by prebuilt NLP rules on the right. Every medical concept in the summary view is linked to an information extraction rule that retrieves a note list which contains relevant information. When users mouseover a summary item, it pops up a window that shows its internal extraction logic and notes retrieved. As seen in the popup window titled ‘NSAIDs’, NLP rules used in DAPR include extensive relevant keyword information and patterns. They greatly reduce search efforts. If users mouseover one of the note list, it drills down to the note content with highlighted keyword information (see the other popup on top of NSAIDs popup). Users can also search for notes and access the contents by user keywords or a regular expression in the left panel’s search box. This figure uses a fictitious patient data for demonstration purpose.

Second, for faster access, we pre-loaded and pre-cached patient data before starting the validation. Currently, DAPR integrates all note data from multiple clinical data resources. Getting patient data takes considerable amount of time and can be a burden to the source systems during working hours. Furthermore, the summary view displays 196 items. Combined with default rules, more than 250 NLP rules needed to be computed to ensure speedy access. Therefore, we pre-loaded all the target patient data during nighttime and computed the NLP rules. The precomputed results are cached in a table so that all the refined information can be loaded immediately. Third, to protect patient privacy and security, we added a module to check an ‘allowed patient list’ for individual validators. We restrict data access to patients assigned to that validator. In addition, we set DAPR up as accessible only by verified users, through our MGB network or VPN. Finally, we built a database and an administrator dashboard to manage users, projects, and audit.

### Validation Process

Fifteen reviewers validated COVID-19 indicators and 22 phenotypes of 150 patients. One experienced reviewer in our group validated 100 patients, and the other fourteen volunteers from the working group validated 55 patients. We divided the 55 patients into 11 groups, 5 patients each. Each validator was assigned at least one data set. The reviewers who were willing to validate more data were given another data set. Twenty seven patients were cross validated. The authors participated as adjudicators for a final decision of any discrepancies between two validators.

We provided data in an encrypted data sheet file. Only the assigned patients were listed in a data file. Data was displayed in one row per patient. We added a row for the validation result and a row for comments below each patient. We also included a data dictionary that described the definition of the data columns, temporal extraction logic, and reasons for inclusion.

Reviewers had freedom to use Epic (i.e., MGB EHR system) and (or) DAPR to validate the assigned data. We required volunteers to receive IRB approval and to take a training about the purpose of the validation and using DAPR for it. Then we assigned patient set(s) and registered them into DAPR. They could access only the assigned patients on DAPR. We also sent a welcome email with instructions and the encrypted patient data table file. A password to open the data file was sent by a separate email. The result was received back by email. All the emails were transferred via secure emails. When the validation is completed, we asked them to participate a survey about the validation experience and the DAPR tool.

## Results

### Validation results

We validated 150 COVID-19 positive patients’ data in the MGB COVID-19 Data Mart. As the validation was a voluntary work, not all reviewers completed all assigned labels. For COVID-19 patient cohort and index date, 102 patients were validated. However, all the other data types were validated for all 150 patients. Total 3,804 labels were reviewed and 697 of them were validated twice by different reviewers.

We evaluated the performance of the selected data types in the COVID-19 Data Mart (Table 2). The COVID-19 positive cohort was highly accurate (95.10% of positive predictive value (PPV)). The data mart mapped COVID-19 related admission correctly at 93.33% PPV, 96.67% negative predictive value (NPV), 95.60% specificity, and 94.92% sensitivity. Extracting the COVID-19 index date (precision 93.14%, recall 100%, and F-measure 96.45%) and admission date (precision 93.33%, recall 94.92%, F-measure 94.12%) also showed high performance.

**Table 2.**
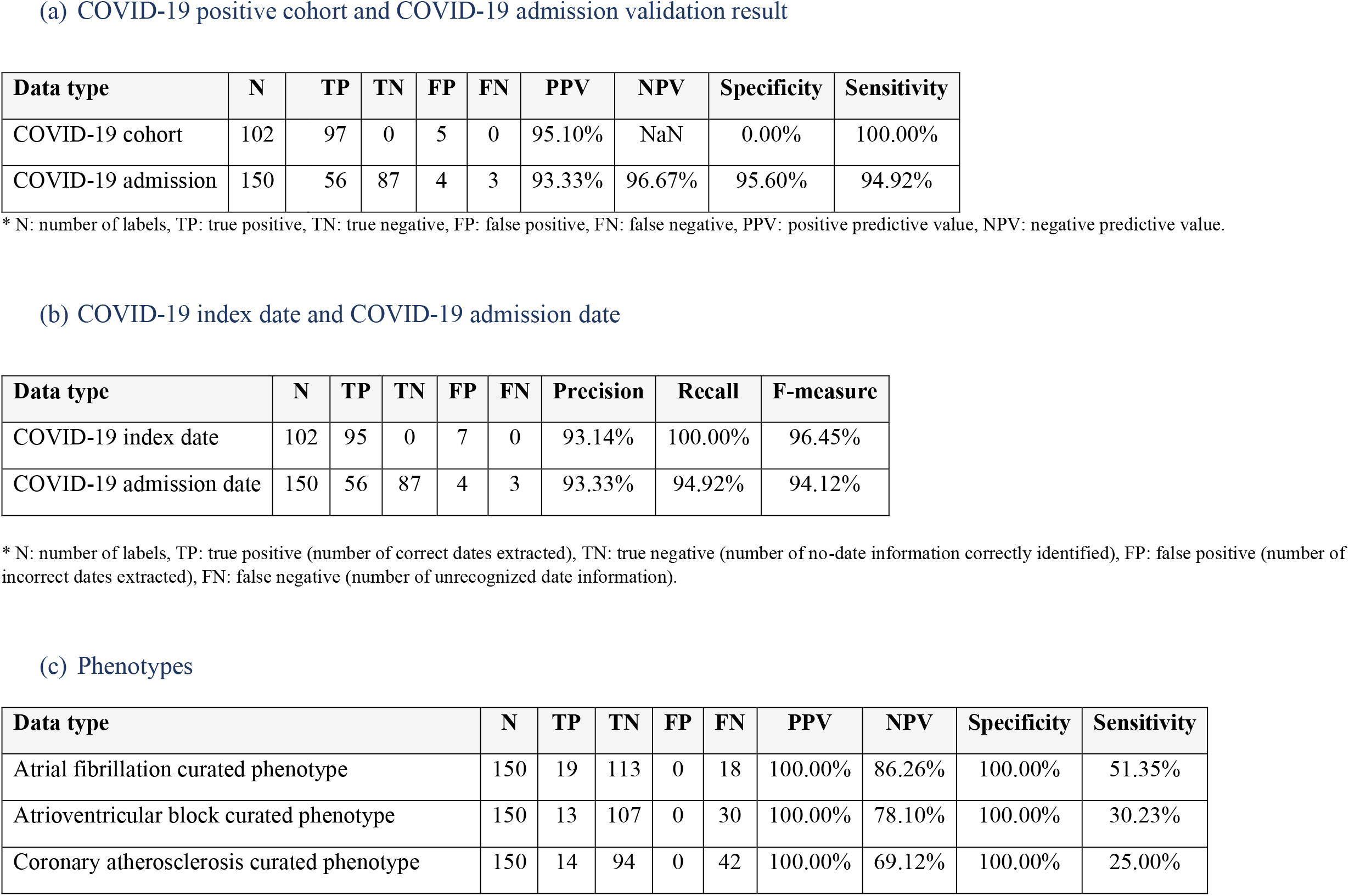
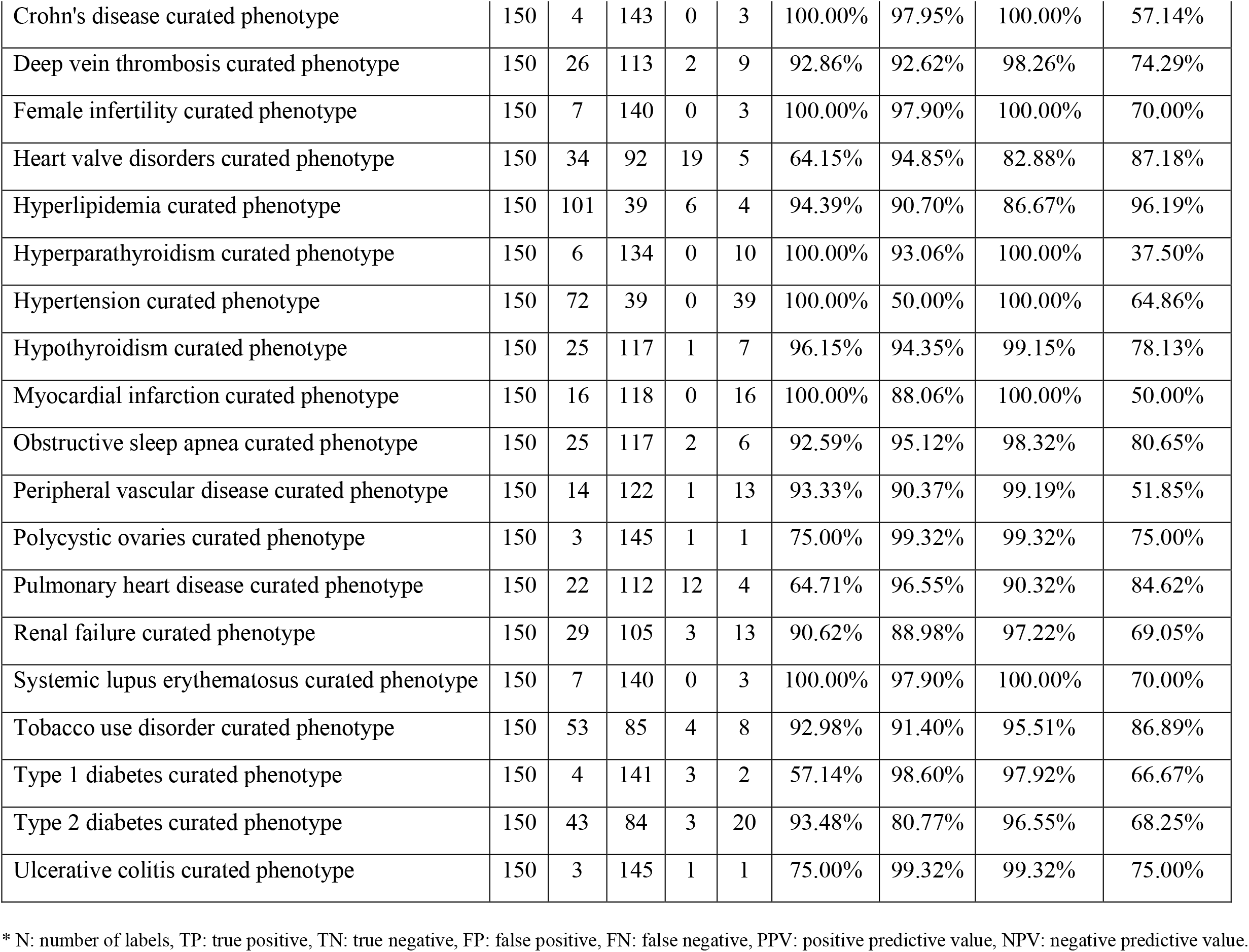
Validation results

(a) COVID-19 positive cohort and COVID-19 admission validation result
| Data type | N | TP | TN | FP | FN | PPV | NPV | Specificity | Sensitivity |
| --- | --- | --- | --- | --- | --- | --- | --- | --- | --- |
| COVID-19 cohort | 102 | 97 | 0 | 5 | 0 | 95.10% | NaN | 0.00% | 100.00% |
| COVID-19 admission | 150 | 56 | 87 | 4 | 3 | 93.33% | 96.67% | 95.60% | 94.92% |

\* N: number of labels, TP: true positive, TN: true negative, FP: false positive, FN: false negative, PPV: positive predictive value, NPV: negative predictive value.

(b) COVID-19 index date and COVID-19 admission date
| Data type | N | TP | TN | FP | FN | Precision | Recall | F-measure |
| --- | --- | --- | --- | --- | --- | --- | --- | --- |
| COVID-19 index date | 102 | 95 | 0 | 7 | 0 | 93.14% | 100.00% | 96.45% |
| COVID-19 admission date | 150 | 56 | 87 | 4 | 3 | 93.33% | 94.92% | 94.12% |

\* N: number of labels, TP: true positive (number of correct dates extracted), TN: true negative (number of no-date information correctly identified), FP: false positive (number of incorrect dates extracted), FN: false negative (number of unrecognized date information).

(c) Phenotypes
| Data type | N | TP | TN | FP | FN | PPV | NPV | Specificity | Sensitivity |
| --- | --- | --- | --- | --- | --- | --- | --- | --- | --- |
| Atrial fibrillation curated phenotype | 150 | 19 | 113 | 0 | 18 | 100.00% | 86.26% | 100.00% | 51.35% |
| Atrioventricular block curated phenotype | 150 | 13 | 107 | 0 | 30 | 100.00% | 78.10% | 100.00% | 30.23% |
| Coronary atherosclerosis curated phenotype | 150 | 14 | 94 | 0 | 42 | 100.00% | 69.12% | 100.00% | 25.00% |
| Crohn's disease curated phenotype | 150 | 4 | 143 | 0 | 3 | 100.00% | 97.95% | 100.00% | 57.14% |
| Deep vein thrombosis curated phenotype | 150 | 26 | 113 | 2 | 9 | 92.86% | 92.62% | 98.26% | 74.29% |
| Female infertility curated phenotype | 150 | 7 | 140 | 0 | 3 | 100.00% | 97.90% | 100.00% | 70.00% |
| Heart valve disorders curated phenotype | 150 | 34 | 92 | 19 | 5 | 64.15% | 94.85% | 82.88% | 87.18% |
| Hyperlipidemia curated phenotype | 150 | 101 | 39 | 6 | 4 | 94.39% | 90.70% | 86.67% | 96.19% |
| Hyperparathyroidism curated phenotype | 150 | 6 | 134 | 0 | 10 | 100.00% | 93.06% | 100.00% | 37.50% |
| Hypertension curated phenotype | 150 | 72 | 39 | 0 | 39 | 100.00% | 50.00% | 100.00% | 64.86% |
| Hypothyroidism curated phenotype | 150 | 25 | 117 | 1 | 7 | 96.15% | 94.35% | 99.15% | 78.13% |
| Myocardial infarction curated phenotype | 150 | 16 | 118 | 0 | 16 | 100.00% | 88.06% | 100.00% | 50.00% |
| Obstructive sleep apnea curated phenotype | 150 | 25 | 117 | 2 | 6 | 92.59% | 95.12% | 98.32% | 80.65% |
| Peripheral vascular disease curated phenotype | 150 | 14 | 122 | 1 | 13 | 93.33% | 90.37% | 99.19% | 51.85% |
| Polycystic ovaries curated phenotype | 150 | 3 | 145 | 1 | 1 | 75.00% | 99.32% | 99.32% | 75.00% |
| Pulmonary heart disease curated phenotype | 150 | 22 | 112 | 12 | 4 | 64.71% | 96.55% | 90.32% | 84.62% |
| Renal failure curated phenotype | 150 | 29 | 105 | 3 | 13 | 90.62% | 88.98% | 97.22% | 69.05% |
| Systemic lupus erythematosus curated phenotype | 150 | 7 | 140 | 0 | 3 | 100.00% | 97.90% | 100.00% | 70.00% |
| Tobacco use disorder curated phenotype | 150 | 53 | 85 | 4 | 8 | 92.98% | 91.40% | 95.51% | 86.89% |
| Type 1 diabetes curated phenotype | 150 | 4 | 141 | 3 | 2 | 57.14% | 98.60% | 97.92% | 66.67% |
| Type 2 diabetes curated phenotype | 150 | 43 | 84 | 3 | 20 | 93.48% | 80.77% | 96.55% | 68.25% |
| Ulcerative colitis curated phenotype | 150 | 3 | 145 | 1 | 1 | 75.00% | 99.32% | 99.32% | 75.00% |

\* N: number of labels, TP: true positive, TN: true negative, FP: false positive, FN: false negative, PPV: positive predictive value, NPV: negative predictive value.

Overall, 22 phenotypes returned good results in PPV (90.11, 95% CI 84.11 –96.11), NPV (89.60, 95% CI 84.44-94.76), and specificity (97.30, 95% CI 95.23 – 99.37). However, sensitivity was measured relatively low (65.90, 95% CI 57.63 – 74.17). Based on this result, 3 phenotypes (*heart valve disorders, pulmonary heart disease, type 1 diabetes curated phenotypes*) which PPVs are less than 70% were removed from the MGB COVID-19 Data Mart and the COVID-19 Summary Table.

### Survey results

Ten validators submitted feedback through the survey. Not all questions were answered by all the survey participants. Most of them used DAPR (i.e., DAPR only or both DAPR and Epic) for the validation, except two participants (Figure 2(a)). DAPR users showed positive attitudes towards DAPR. They commented DAPR is great, helpful, and easy to use. Especially, the summary view was mentioned as useful. P5 said *“I much preferred using DAPR over Epic. It seems to search notes that I would never find on Epic, or that would take far longer to do so. … DAPR seems more helpful to me for specific phenotype searches*.*”*

**Figure 2.**
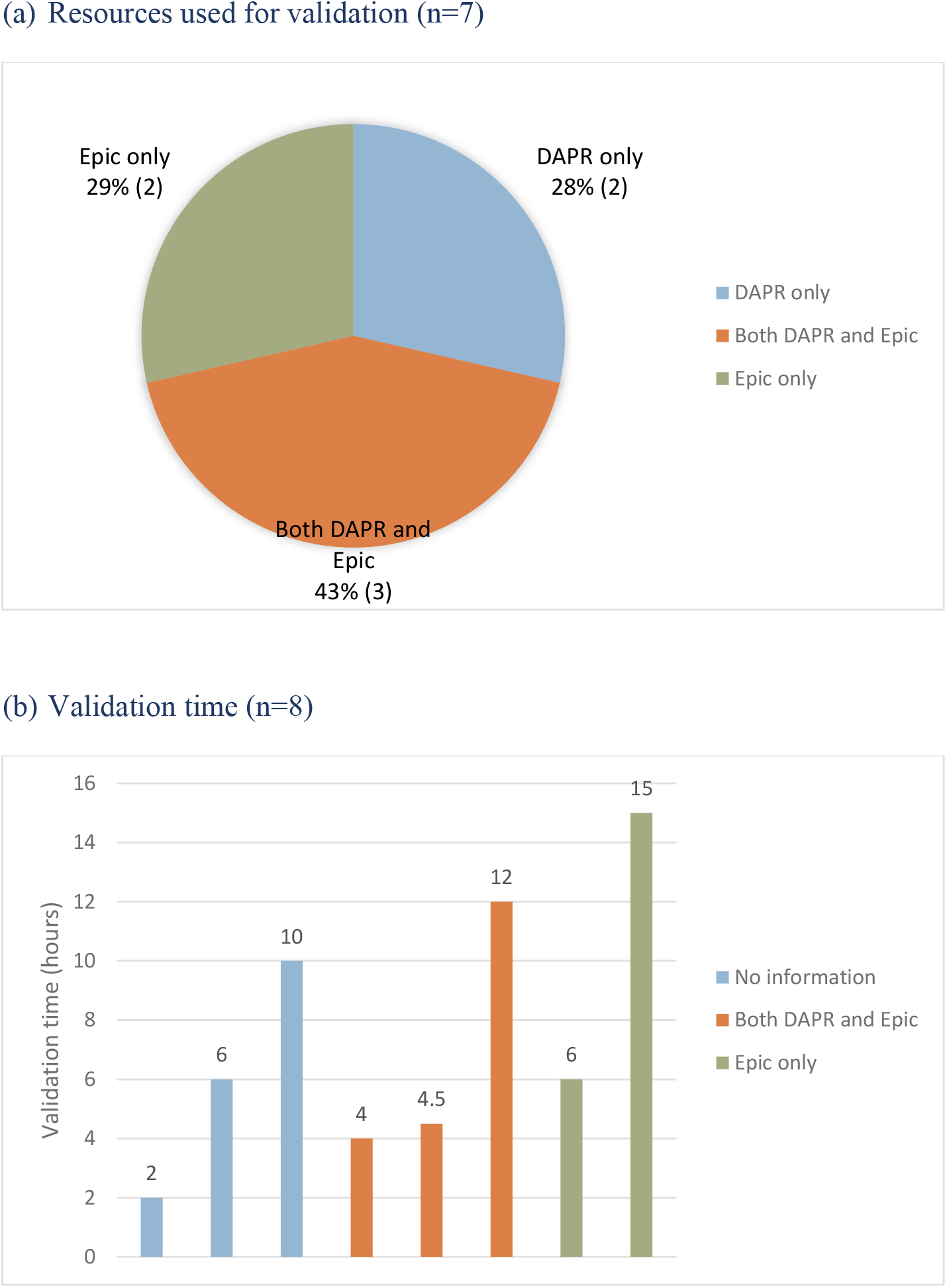
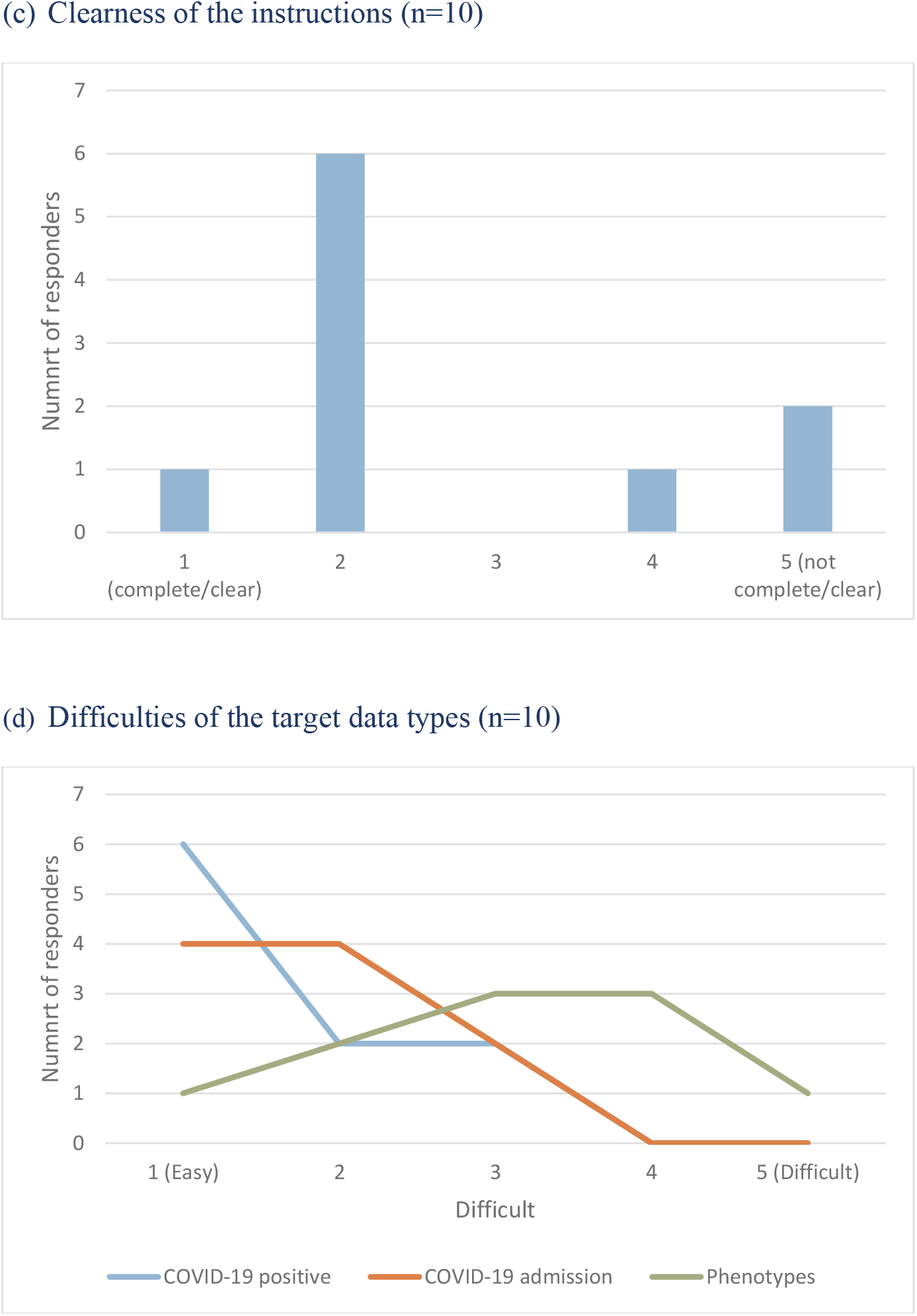
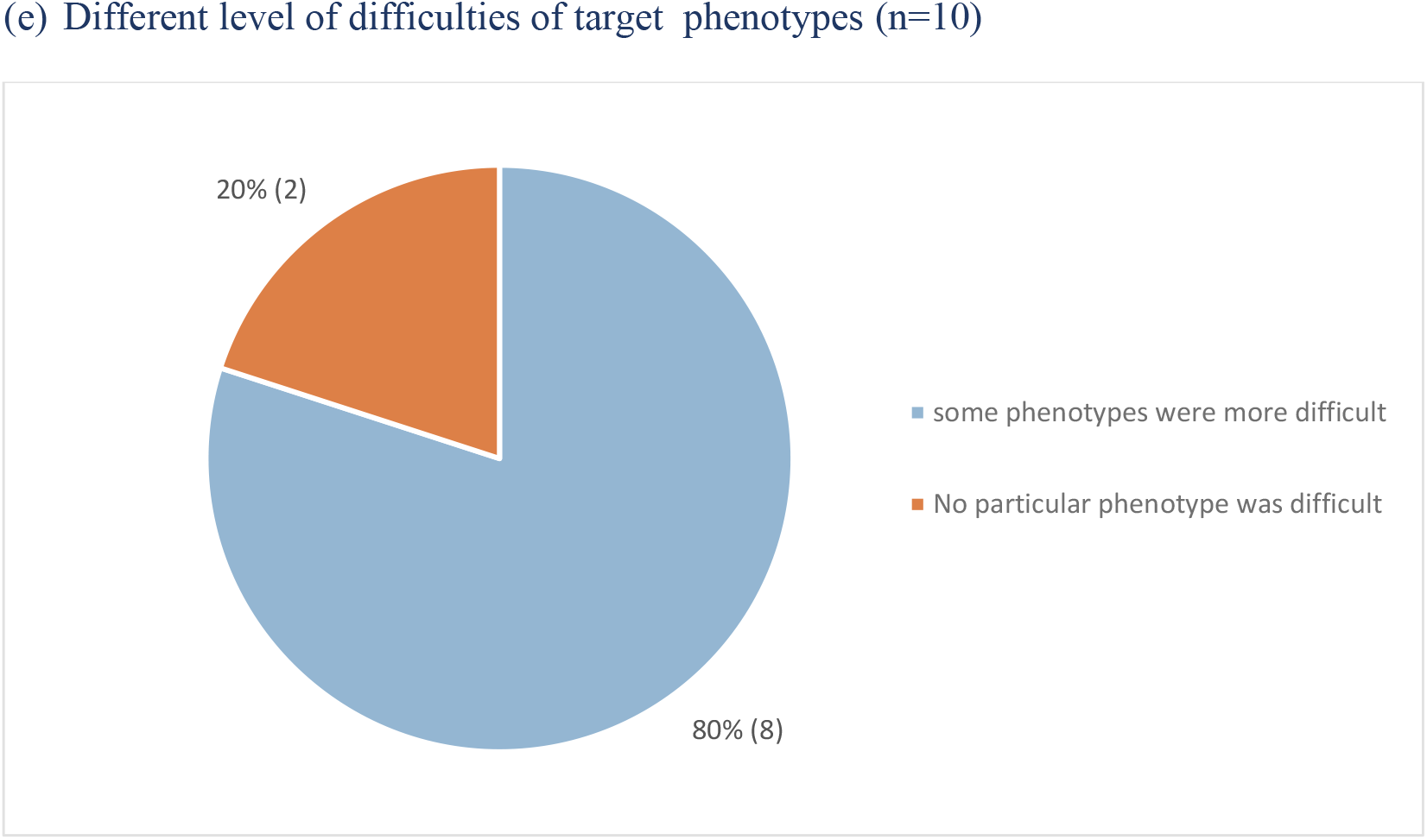
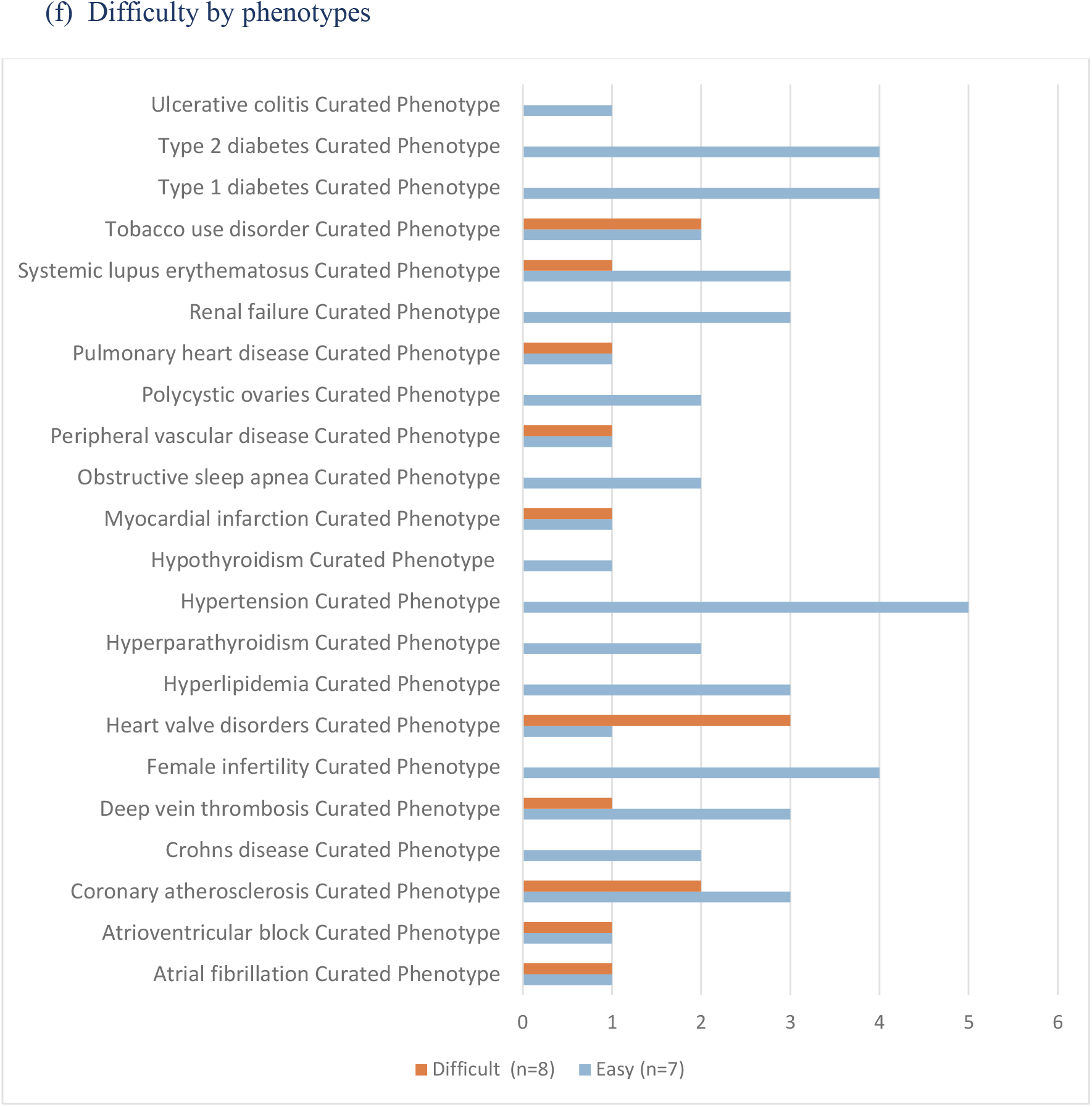
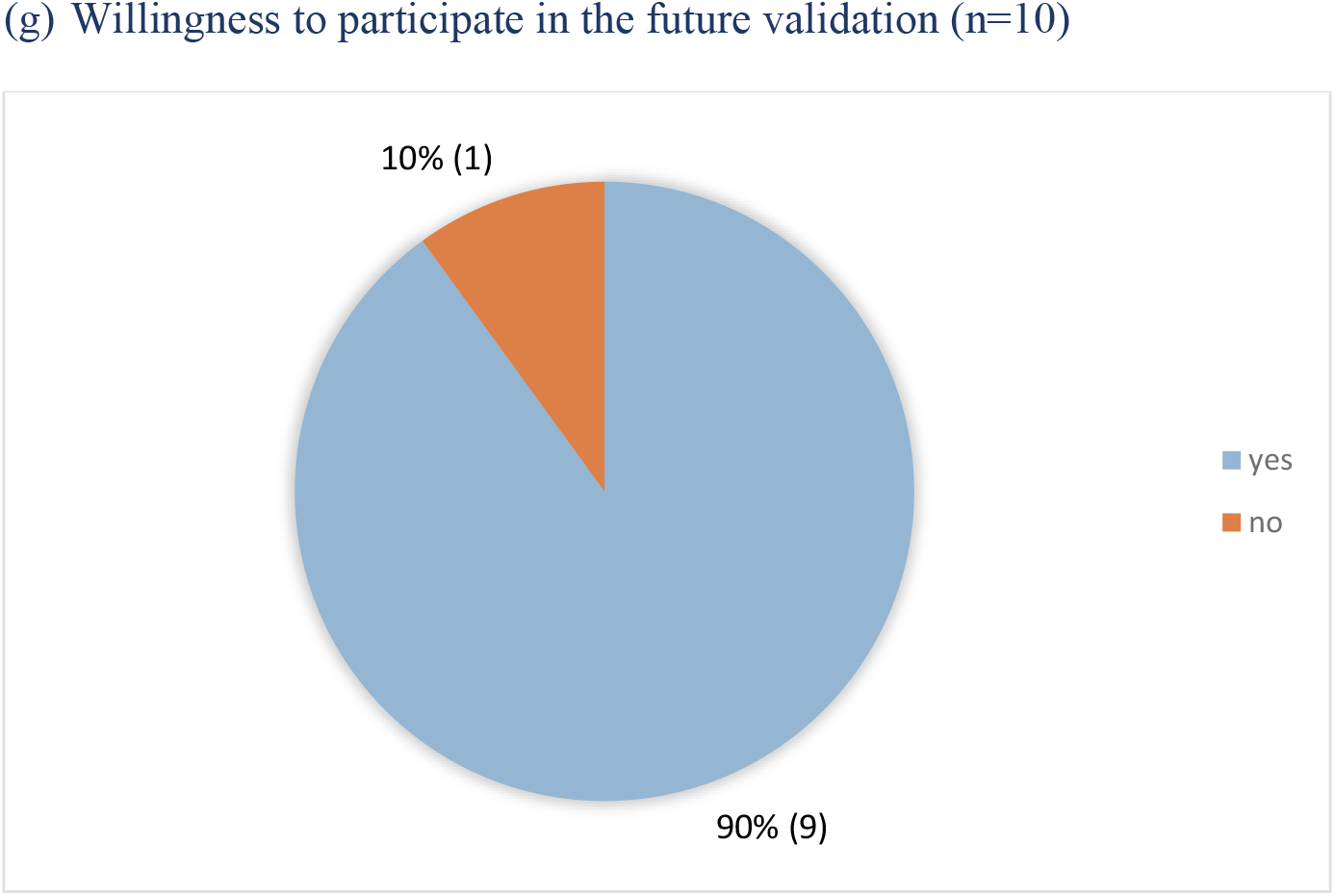
Quantitative survey results (N=10) This figure illustrates quantitative survey results only. Qualitative results are summarized in the manuscript.

However, P5 asserted using DAPR and EPIC in tandem was the most helpful, since Epic is great for getting an overall snapshot of the patient’s history. This aspect is linked to a suggestion on DAPR, by another participant (P7), to have a visualization of events along time that directs users to records. Other ideas such as improving summary view performance (P2, P7), displaying performance values for curated phenotypes (P2), and removing redundant phenotypes (P7) in the summary view were also submitted to improve DAPR. Conversely, there were negative experiences reported. Difficulty caused by mouse out interaction was pointed as a drawback: *“mouseover was challenging-I wanted to keep a pdf preview window open without losing a note date. I had to retrace my steps over a dozen times”* (P4). Other poor experiences occurred by misuse (P4) or a temporary issue (P6) were received.

Participants expressed the validation work was straightforward and mostly easy to complete. P5 stated *“I found this process to be very straightforward and, other than some of the phenotypes, easy to complete*.*”* The time spent for validation ranged from 2 hours to 15 hours. The validators who informed that used both DAPR and Epic spent 4 ∼12 hours and the ones used Epic alone took 6 ∼ 15 hours (Figure 2(b)). Instruction materials were deemed clear and helpful for most of the participants (Figure 2(c)).

There were suggestions to improve instructional materials. Having an extra field to mark certainty, providing clearer phenotype definitions, and giving more guidelines for decision strategies were proposed. P2 suggested *“A column or field to let the validator input the certainty of the agreement/disagreement pieces of evidence may have some value in evaluating the results*.*”* P8 pointed that *“The definition of some of the comorbidities is not very specific*.*”* Regarding instructions, P6 said they were not clear *“whether conditions need to be active or just need to be in the history”*. Furthermore, R1 shared *“I had a difficult time determining what notes/lab results/comments were required to make a phenotype go to ‘YES” so more instructions on this for someone with limited clinical experience would be helpful*.*”* These are related to difficulties the reviewers experienced in some phenotypes.

Eighty percent of the respondents agreed on *‘some phenotypes were more difficult to validate’* while no one answered validating COVID-19 positive and COVID-19 admission was difficult (Figure 2(d) and (e)). *Heart valve disorders* was mentioned as difficult by most validators, followed by *Coronary atherosclerosis* and *Tobacco use disorder*. In contrast, *Hypertension, Female infertility, Type 1 diabetes*, and *Type 2 diabetes* were the top two ranked phenotypes easy to validate (Figure 2(f)).

Participants explained the difficulty was caused by lack of medical knowledge, confusing phenotype definitions, and unclear decision strategy. Even though DAPR helps note search by prebuilt NLP rules, some phenotypes still require users’ medical knowledge. For example, if there is no definitive evidence from a phenotype’s result, validators need to choose many data factors for further investigation to make decisions. P1 said *“The phonotypes I found difficult seemed to have many different words for same thing, the easier ones seemed to always use the same terminology*.*”* P7 added a detail of the same context, *“I, being lack of clinical training, it took more time connecting diseases, signs, tests, medication etc. together to make a decision*.*”* P5, who noted is an experienced reviewer in the survey comment, also had difficulties in certain phenotypes due to lack of specialty in that domain: *“Those particular phenotypes were harder to discern because there seemed to be many ways they could be listed or noted in the patient’s chart. It was harder for me, since I’m (obviously) not a cardiologist, to be able to clearly separate these diagnoses from others*.*”*

In addition, the extraction coverage being different from its phenotype definition or redundant phenotype items returning different list arose confusion to reviewers. These made them spend extra efforts to understand how the phenotype is identified. Although P2 expressed *“I feel that the current summary table puts the sensitivity into a higher priority compared with specificity. This is reasonable and maybe the best choice*.*”* Many pointed out that the returned note list do not exactly support the (non-)existence of the phenotypes: *“[The prebuilt logics] might not have captured the whole situation for a phenotype”* (P7), *“the definition and the query algorithm seem to be different”* (P8), *and “[Summary items return] more evidence and related diseases in that category”* (P2). Moreover, P1 mentioned about the redundant summary view items, *“Sometimes the past medical history did not seem to match the phenotypes. For example, there are two valve diseases, one in past medical history section and the other in phonotype section which were not always consistent”*

Consequently, participants had questions about the phenotype definitions: *“The definition/scope of some of the phenotype was ambiguous/wide”* (P3), “*not clear definition of a phenotype to me*” (p7). They shared that they had to go through multiple notes (P9: *“Had to go through multiple notes to look for the info”*) or cross patient comparison (R7: *“cross patients comparison was also done to understand why one patient is Y and the other is N for a specific phenotype*.*”*) to understand the phenotype definitions.

Lastly, participants were uncertain about the validation strategy in general for some specific conditions. P9 was not sure whether past phenotype should be marked Y or N: *“Determine if condition still active - if past condition, not sure if that should have been yes or no. For example, PAF or renal failure that resolved but patient was on dialysis temporarily”*. P10 said *“some more direction on additional rarer conditions that might impact the data [is needed]*.*”* For example, P4 commented that *“The relative sensitivity required was unclear. If there was a single instance of a drug administered for DVT 10 years ago (among hundreds of possible notes) and no other terms or diagnoses, should that person be coded as having DVT?*.*”*

Participants showed satisfaction for being involved in this validation work: *“Thanks for letting me in this project”* (P7) and *“delighted to be involved”* (P10). Nine participants showed willingness to participate in the future validation again (Figure 2(g)).

## Discussion

One of the biggest problems of validating COVID-19 data was that large number of COVID-19 patients do not have rich data in our system. Many of them were transferred patients or were previously healthy. This situation created the need for note review to discover patient history not existing in coded data fields. DAPR provided a way to use NLP in an automated way to extract buried patient information. In the survey, a participant noted using DAPR was more useful than just searching notes via Epic due to its extensive search ability.

In addition to reviving the QPID service as DAPR, we made efforts to repurpose it to support COVID-19 validation. We created a custom COVID-19 validation summary view and developed information extraction logics through MGB RISC NLP pipeline. We also made a module to facilitate testing and uploading the NLP rules efficiently. Moreover, we considered practical and proper use of the tool. To enable faster access, to protect patient privacy and security, and to manage administrative issues, we built formal processes and additional system modules such as precaching, checking allow list, administration tool, etc.

Based on the survey results, the main barriers that created difficulty for some phenotypes were caused by the NLP rules’ coverage and wide definition of the phenotypes. While one said prioritizing sensitivity over specificity is reasonable and maybe the best choice, many pointed the extraction results not exactly describing the phenotype definition made them feel difficult. For example, when there is no definitive evidence, one should take extra efforts to explore multiple data factors to find the clues. Moreover, although the summary view’s logic not only reduce search efforts and help a user’s lack of specialty by built in relevant keywords, it still required users’ background knowledge to select which ones to investigate.

In addition, redundant summary items were returning different notes added difficulty. When we added the *PHENOTYPES* category in the DAPR summary view, there were two phenotypes that remained appear in other categories (*hypertension* in *COVID-19*’s *Risk Factors* and *valve disease* in *Select Past Medical History*’s *CV, which section is for cardio vascular diseases*) due to the importance of the data in that context. We assigned different rules for the curated phenotypes, reflecting the logics in the phenotyping algorithms. Consequently, DAPR produces different results for the same named disease.

Some participants commented the phenotype definitions and decision guidelines in the instructions were wide and ambiguous. This is a very different situation compared to other phenotype validations have been done in our group. Typically, a validation is focused on a narrow domain, and few trained experts participated and handled the decision strategy. The phenotypes selected for the COVID-19 data validation are the ones that had been validated years ago. At that time, we did not have to provide further details than what we currently described. However, for this validation, everyone had to deal with broad scope of domains, which might not be familiar with their specialty, in a short amount time. The comments confirm the challenges of the COVID-19 validation.

Lower sensitivities found in phenotypes could be attributed to the timing of the last algorithm run. We ran the phenotyping algorithms in March 2020, but the validation was started at the end of June 2020. Therefore, if there were more data added after the phenotyping ran, it could have affected the sensitivity. Another possible explanation is incompleteness of the COVID-19 patients’ data. Since significant number of COVID-19 patients are new patients, there might not have been enough data to conclude a phenotype algorithmically. However, further studies are needed to confirm.

One participant showed interest of using DAPR in their own research, in the survey. COVID-19 is not the only domain that could benefit from DAPR. Many data in the summary view are commonly used indicators in patient review that are useful. There have been already requests to use DAPR from multiple groups. Several validation projects are underway using DAPR. On the other hand, currently, some workflow steps remain manual and limit wide use. We are working to automate and operationalize it to meet growing demands.

## Conclusion

In this study, we used an NLP-based chart review tool, DAPR, for COVID-19 associated data validation and contributed building a reliable COVID-19 research data mart. We transitioned DAPR from a clinical tool to a research tool. We designed a COVID-19 relevant patient summary view and built new information extraction rules. We enabled faster loading by preloading and precaching the patients. We added components to safeguard patient privacy, to harden information security, and to provide auditing capabilities to adhere to IRB governance. We also designed a new workflow to use DAPR for validation.

Fifteen reviewers validated COVID-19 indicators and 22 phenotypes of 150 patients in the MGB COVID-19 Data Mart, using either or both DAPR or (and) Epic. The overall statistical results (PPV, NPV, sensitivity, and specificity) showed good performance in all data types, except sensitivity of phenotypes. As a result, three lowest PPV phenotypes were removed from the COVID-19 Data Mart and the COVID-19 Summary Table. The participants thought DAPR is easy to use and facilitates the validation work. Especially, DAPR’s summary view eased users finding relevant information.

The results show how the use of NLP technique can help overcome unusual challenges brought by COVID-19. Although the reviewers had various clinical backgrounds and they had to find information for wide domains of target data mostly from notes, all of them could complete the tasks in a short amount of time. However, in some phenotypes, the outcomes extracted by NLP rules were unable to capture all possible situations. It required users to make additional efforts to search for clues using their knowledge. It remains as a limitation of using an NLP tool for validation.

DAPR is heavily NLP-driven. Its NLP rules are highly customizable and generalizable. They can be tailored to search notes of differing formats and templates from different institutional sources to give reviewers a unified view. In this case study, while we have demonstrated the applicability of DAPR to notes from MGB institutions, we think the DAPR’s approach for note reviews is generalizable to other institutions given sufficient customization of the NLP rules.

For the next phase, we will validate COVID-19 signs and symptoms in the data mart. Moreover, we plan to operationalize DAPR as a pilot service for wider MGB research groups. Currently, there are steps that require manual interventions to initiate a new project on DAPR. We are in the process of automating the DAPR use workflow.

## Data Availability

Due to privacy concerns, the source data cannot be made openly available.

## Acknowledgments

This study was approved by the Institutional Review Board (IRB) of Mass General Brigham under protocols *Digital Analytic Patient Reviewer (DAPR) (2020P000605)* and *Validating EHR data in the COVID-19 Mart and Summary Table (2020P001639)*. The authors thank all the validators and attendees of the MGB Center for COVID Innovation Clinical Trial Tools & MGB COVID-19 Data Mart Working Group for the valuable validation work and comments.

